# Smartphone applications in the prevention of Type 2 Diabetes Mellitus: a protocol for a systematic review

**DOI:** 10.1101/2020.05.18.20106211

**Authors:** Esrat Jahan, Rawan Almansour, Kiran Ijaz, Rimante Ronto, Liliana Laranjo

**Affiliations:** Faculty of Medicine and Health Sciences, Macquarie University; College of Applied Medical Sciences, King Faisal University; Australian Institute of Health Innovation, Macquarie University

## Abstract

Diabetes mellitus is a leading cause of concern among non-communicable diseases worldwide, with its prevalence increasing every day. Studies have shown that it is possible to prevent type 2 diabetes in high risk people if they adopt a healthy lifestyle, such as exercising regularly, eating nutritious food and maintaining an ideal weight. Mobile apps may aid these people in maintaining a healthy lifestyle. Till date, no systematic review has evaluated the use of mobile health applications for the prevention of type 2 diabetes. In this systematic review protocol we will lay out the methods we will use to synthesise the evidence about mobile health applications for the prevention of type 2 diabetes, focusing particularly on their impact on different process and outcome measures, as well as on patient perspectives. Database searches will be done in PubMed, Embase, CINAHL and PsychInfo. Screening of the articles will be conducted by two independent researchers. The Cochrane risk of bias tool will be used for quality assessment. A narrative synthesis of the included articles will be done and the results summarised. The findings of this review will provide evidence on the impact of mobile applications in preventing Type 2 diabetes mellitus.

## Background

Diabetes has been labelled the ‘biggest epidemic of the twenty-first century’. (1) The number of people being diagnosed with diabetes has been increasing, with an estimated 463 million people suffering from the disease worldwide (1, 2) The figures are predicted to rise to about 578 million by 2030 and 700 million people by the year 2045, with an increase in prevalence from 8.8% to 9.9%. (1, 3) The mortality rate for diabetes is also high, causing 1.6 million deaths and making it the seventh most common cause of death worldwide. (4) Complications related to diabetes, such as cardiac diseases and stroke also contribute to this rate. (4) There are mainly two types of diabetes, type 1 and type 2, with type 2 diabetes constituting around 95% of all diabetes cases and contributing more significantly to the disease burden. (5, 6) According to a national health survey conducted by the Australian Bureau of Statistics, about 1.2 million Australian adults were suffering with the disease in the year 2017-2018, costing the government $2.7 billion dollars in managing it. (7, 8)

Type 2 diabetes is chronic in nature, in which the body cannot effectively use the insulin that is being made by the pancreas. (9) Insulin helps in the metabolism and regulation of glucose levels. People with diabetes have increased blood glucose levels. (10) Type 2 diabetes is mainly caused by poor lifestyle choices, such as unhealthy eating, inadequate physical activity, lack of sleep, excessive consumption of alcohol and smoking. (11) Studies conducted in various parts of the world have shown that it is possible to reduce the incidence of Type 2 diabetes for people at risk of developing the disease, via lifestyle modifications such as reduction of body weight, increase in physical activity and improving dietary intake. (12, 13)

Interventions aimed at preventing diabetes are usually provided in a community setting approach, where the group of individuals and health care workers (doctors, nurses, dieticians, community healthcare workers) meet together, at certain intervals for a specific period, to bring about desired changes in lifestyle habits. (14, 15, 16) Most of these strategies are aimed at people who are clinically classified as “prediabetes”, i.e. people whose blood glucose levels fall between the ranges of normal values and the values of Type 2 diabetes. (14) However, given that about 352 million adults fall under this category, which is around 7.3% of the population, there is a need for interventions that can be easily disseminated at scale for these groups of people.

In recent years, mobile health technologies have been employed to deliver health care interventions, especially in the prevention and management of chronic diseases. (17) Previous studies focusing on text messaging have shown promising results in motivating people to take up an active lifestyle to prevent Type 2 diabetes. (18, 19, 20, 21, 22) More recent mobile technologies, such as smartphone applications, have the potential to offer more engaging interventions, available at any time, when patients may need them most. Half of the world’s population own a smartphone, with a recent report stating that about an estimated 3.5 billion people have one. (23) Additionally, 70% of people living in developing nations are mobile phone users, where the disease burden for diabetes is increasing. (24, 25) So far, no systematic review has evaluated the role of smartphone or tablet applications in preventing diabetes.

## Objectives

The aim of this systematic literature review is to synthesize primary studies focusing on the use of mobile health applications for the prevention of type 2 diabetes, particularly their impact on different process and outcome measures, as well as patient perspectives.

## Methods

### Database search

Articles will be searched in the electronic databases Pubmed, Embase, CINAHL and Psychlnfo. The search terms will include “mobile phone OR cell phone OR smartphone”, “app OR application OR mobile application”, “type 2 diabetes OR diabetes mellitus” AND “prevention”, “prediabetes OR impaired glucose tolerance”(complete search strategy is provided in Supplement 1). The search will be limited to human studies and articles published after 2008, when the first app stores were launched. The reference lists of relevant articles and grey literature such as dissertations, theses, and conference proceedings will be screened to ensure all eligible studies are included for analysis. We will contact the authors to retrieve any additional information about the studies if necessary.

### Eligibility criteria

The inclusion and exclusion criteria set for this review, according to the PICO framework, are:

#### Population

Studies focusing on adults (18 years and above, up to 65 years) at risk of developing type 2 diabetes (e.g. such as physically inactive, obese), as defined by the authors of the studies. Studies will be excluded if they recruit both prediabetes and Type 2 diabetes mellitus participants, where the results are not shown separately for each group.

#### Intervention

We will include any intervention involving a mobile device application in a smartphone or tablet, designed specifically for patients at high risk of developing type 2 diabetes to adopt a healthy lifestyle (e.g. maintaining ideal weight, being physically active, and/or consuming healthy food), with the aim of preventing type 2 diabetes. We will exclude interventions focusing exclusively on other mobile health technologies (e.g. text messaging, telehealth) or internet and web-based interventions.

#### Comparator

Any comparisons for experimental studies will be included. Studies without any control group (e.g. qualitative studies) will also be included, as patients’ perspectives will also be examined.

#### Outcome

Any outcomes related to diabetes [e.g. glycated hemoglobin (HbA1c), glycemia, % participants developing type 2 diabetes], weight, body mass index, and changes in lifestyle behaviours (e.g. eating habits, level of physical activity) will be included. Articles exploring patient perspectives regarding the use of the intervention will also be added.

#### Study designs

Experimental studies (RCTs and quasi-experimental with 1 or 2 arms), qualitative studies exploring patient perspectives on using a mobile app to prevent type 2 diabetes, or mixed methods studies (experimental + qualitative) will be included. Conference abstracts will be excluded.

### Screening

The references retrieved from the databases will be exported and stored in EndNote and the initial screening will be done in Rayyan. Two independent investigators will conduct the initial screening of studies based on the information in their titles and abstracts and then will perform a full-paper screening. Any disagreements will be solved through discussions with a third investigator. Cohen’s kappa will be used to measure inter-coder agreement.

### Data extraction and quality assessment

One reviewer will extract information from the included studies into an electronic extraction form. Two investigators will review the complete extraction form for consistency. The data that will be extracted from the study will include: author, year of publication, country, study design, sample size, population, mean age of population, time of follow-up, intervention characteristics, comparison, and outcomes. The complete eligible studies will be reviewed by two investigators to appraise their quality (using the Cochrane risk of bias tool, when appropriate), and disagreements will be resolved by consensus.

### Synthesis of results

A narrative synthesis will be conducted for all the included studies. The Preferred Reporting Items for Systematic Reviews and Meta-Analyses reporting (PRISMA) statement (26) will be followed when conducting this review, synthesising the results and writing the final manuscript.

## Data Availability

As this is a protocol paper, no data will be available.

